# Human behaviour, NPI and mobility reduction effects on COVID-19 transmission in different countries of the world

**DOI:** 10.1101/2022.04.29.22274485

**Authors:** Zahra Mohammadi, Monica Gabriela Cojocaru, Edward Wolfgang Thommes

**Author notes:** Equal contributor.

## Abstract

**Background:** The outbreak of Coronavirus disease, which originated in Wuhan, China in 2019, has affected the lives of billions of people globally. Throughout 2020, the reproduction number of COVID-19 was widely used by decision-makers to explain their strategies to control the pandemic.

**Methods:** In this work, we deduce and analyze both initial and effective reproduction numbers for 12 diverse world regions between February and December of 2020. We consider mobility reductions, mask wearing and compliance with masks, mask efficacy values alongside other non-pharmaceutical interventions (NPIs) in each region to get further insights in how each of the above factored into each region’s SARS-COV-2 transmission dynamic.

**Results:** We quantify in each region the following reductions in the observed effective reproduction numbers of the pandemic: i) reduction due to decrease in mobility (as captured in Google mobility reports); ii) reduction due to mask wearing and mask compliance; iii) reduction due to other NPI’s, over and above the ones identified in i) and ii).

**Conclusion:** In most cases mobility reduction coming from nationwide lockdown measures has helped stave off the initial wave in countries who took these types of measures. Beyond the first waves, mask mandates and compliance, together with social-distancing measures (which we refer to as *other NPI’s*) have allowed some control of subsequent disease spread. The methodology we propose here is novel and can be applied to other respiratory diseases such as influenza or RSV.

## 1 Background

The first known case of disease caused by SARS-CoV-2 was identified in Wuhan, China, in December 2019. The disease spread worldwide in a few weeks, leading to a pandemic still ongoing as of the winter of 2022. Since December 2019, the basic and effective reproduction number of COVID-19 have been continuously discussed by scientists, political decision-makers and the media (regular and social). The basic reproduction number of an infectious disease, denoted by *R*_0_, represents the expected number of new cases generated by a single infectious individual in a fully susceptible population. In epidemiology, a pandemic will be under control and the transmission will die out when *R*_0_ *<* 1. Estimating the basic reproduction number for COVID-19 was challenging since it was depending on the behavioural activity in local populations. The basic reproduction number for COVID-19 was reported to take values from 2.2 [1] to 6.33 [2] in different countries. In general, although whole populations started with being susceptible to virus, the progression of the pandemic steadily decreased the number of susceptibles in each region. Consequently, the average number of secondary cases per infectious case changed as the population became immunized (by recovering or dying). Thus as the pandemic progressed, *R*_0_ gives way to the effective reproductive number, denoted by *R*_*eff*_, which measures the average number of secondary cases per infectious case in a population at any specific time (where only a fraction is susceptible) [3]. In epidemiology, *R*_*eff*_ is a monitoring indicator of progress in controlling a pandemic. It can also be a way to monitor the effectiveness of interventions (both the effect of immunity and of additional non-pharmaceutical interventions) during a pandemic.

Transmission of SARS-COV-2 depends on the rate of person-to-person contact and on the probability of transmission given one meaningful contact between an infected and a susceptible individual. While the probability of transmission per contact is reduced by NPI’s, principally the wearing of masks [4, 5], the effectiveness of mask-wearing in preventing transmission of SARS-CoV-2 was not recognized in some world regions during the first several months of the pandemic, despite the effectiveness of public mask use in controlling the spread of the 2003 SARS [6, 7].

Another critical non-pharmaceutical intervention for slowing epidemic growth is social distancing which includes shelter-in-place requirements, prohibition of indoor gatherings, imposition of travel restrictions, school closures and workplace closures. Previous studies investigated the quantitative relationships between the COVID-19 epidemic parameters (for instance the total death toll) and social distancing efforts [8, 9, 10, 11]. Mobile data provide a unique opportunity to investigate to some degree the effectiveness of social distancing measures in reducing the effective reproduction number of COVID-19 [12, 13, 14]. Mobile data can be interpreted as a proxy of person-to-person contact reduction as a consequence of social distancing measures, although it generally does not capture short-range behavioral changes, such as maintaining a 6 foot spacing between individuals in public settings. The publicly available data on human mobility that is provided by Google, Apple, Facebook, etc have been used in several articles to evaluate the effectiveness of non-pharmaceutical interventions (NPIs) on the spread of COVID-19 [15, 16, 17, 18, 19, 20].

Existing pre-pandemic work, such as [21, 22, 23], represents the effect of the population-level contact patterns on the infection transmission dynamics. The infection transmission rate varies in different countries as a result of the different social and economic structures and different contact patterns. Mistry et al [22] used the derived contact matrices to model the spread of airborne infectious diseases. The transmission rate of COVID-19 fundamentally depends on interpersonal interaction rates. Most countries considered different strategies for reducing the contact rate in order to control the spread of the virus. Feehan and Mahmud calculated age-structured contact matrices to quantify how much interpersonal contact has changed during the pandemic in the United States [24]. They estimated about 82% decline in interpersonal contact between March 22nd and April 8-th, 2020 (known as wave 0) and an increase in daily average contact rates over the subsequent waves. Other studies observed the decline in contact rates in China [25], United Kingdom [26], Luxembourg [27], Italy, Belgium, France, and the Netherlands [28] throughout the pandemic. Prem et al. created synthetic contact matrices to represent the effect of intervention measures to reduce social mixing on outcomes of the COVID-19 epidemic in Wuhan, China [29, 30]. The age-specific social contact characterization also supports the possibility of suspecting differences in transmission patterns of COVID-19 outbreak among different age-groups [31, 32, 33, 20]. In this work, projected contact matrices provided by Prem et al. [21] are used.

This work presents a SEIRL (Susceptible, Exposed, Infectious, Recovered and isoLated) model that uses incidence data, Google mobility data (as a modifier of effective contact rates), mask efficacy and mask compliance data to provide insight into the strategies employed for controlling the pandemic in each country under consideration. It explores and quantifies the effectiveness of non-pharmaceutical policy interventions across 12 diverse countries/regions, using their effective reproduction number *R*_*eff*_. To accomplish these goals we estimate the *R*_0_ values of each region (using the classic next-generation matrix analysis [34]) and then a time-dependent effective reproduction number, denoted by *R*_*eff*−*data*_, using known repositories of incidence data for each country [35]. Using the projected contact rates from [21] extrapolated to each country of interest with publicly available demographic data, we also infer the time-dependent effective contact rates, and hence the time-dependent effective reproduction number of each country as a function of multiple Google mobility indices. We denote this by *R*_*eff* −*mobil*_.

Comparing the two estimates of *R*_*eff*_ we investigate the effect of NPI’s, over and above changes in contact rates due to mobility and mask mandates, in each country’s epidemiology throughout the year 2020^[1]^. It is concluded in most cases that mobility reduction coming from stringent lockdown measures helped stave off the initial wave in countries which took these types of measures (see for instance Figure 2). While mobility increased in the second part of 2020 (see Figure 7), mask mandates together with all other NPI measures (for instance social-distancing) have allowed some “control” of subsequent waves, in the sense that countries were able to maintain their effective reproduction numbers around 1.

**Figure 1:**
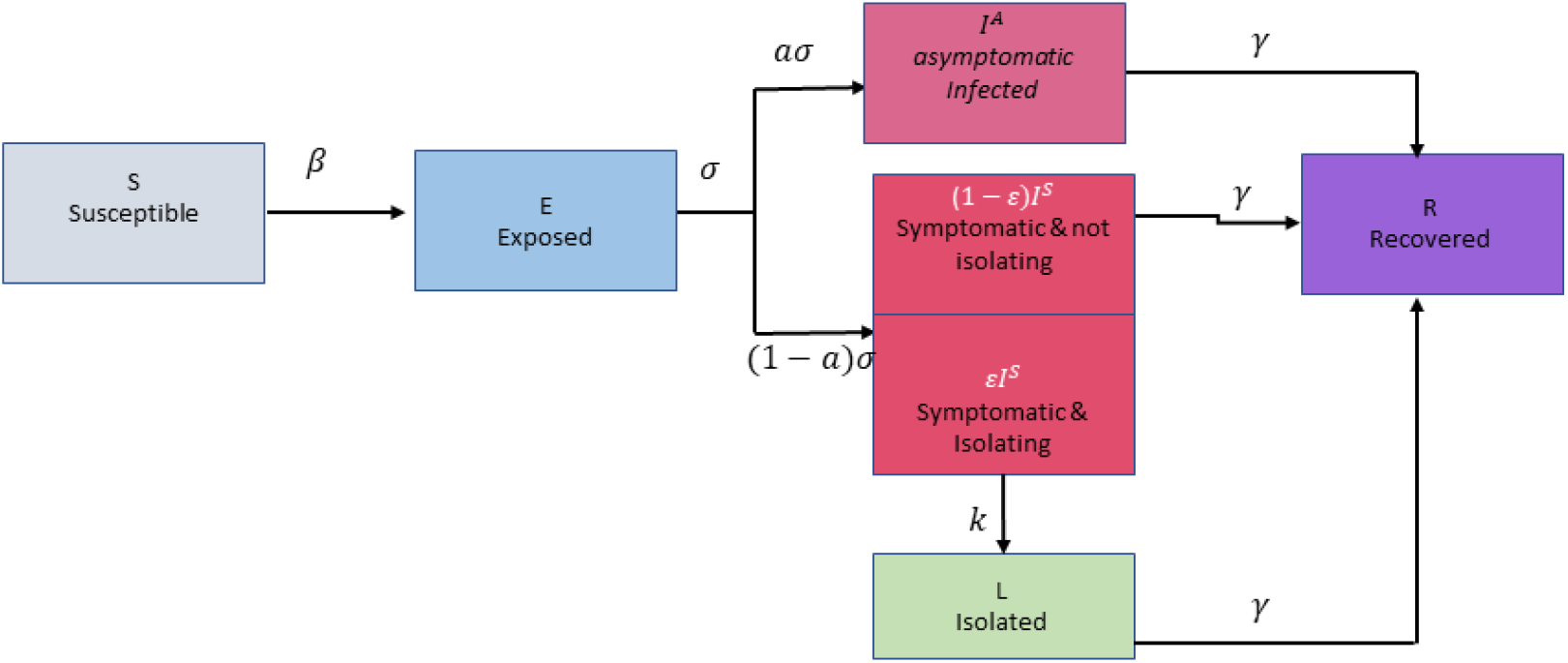
Transmission model in diagram form.

**Figure 2:**
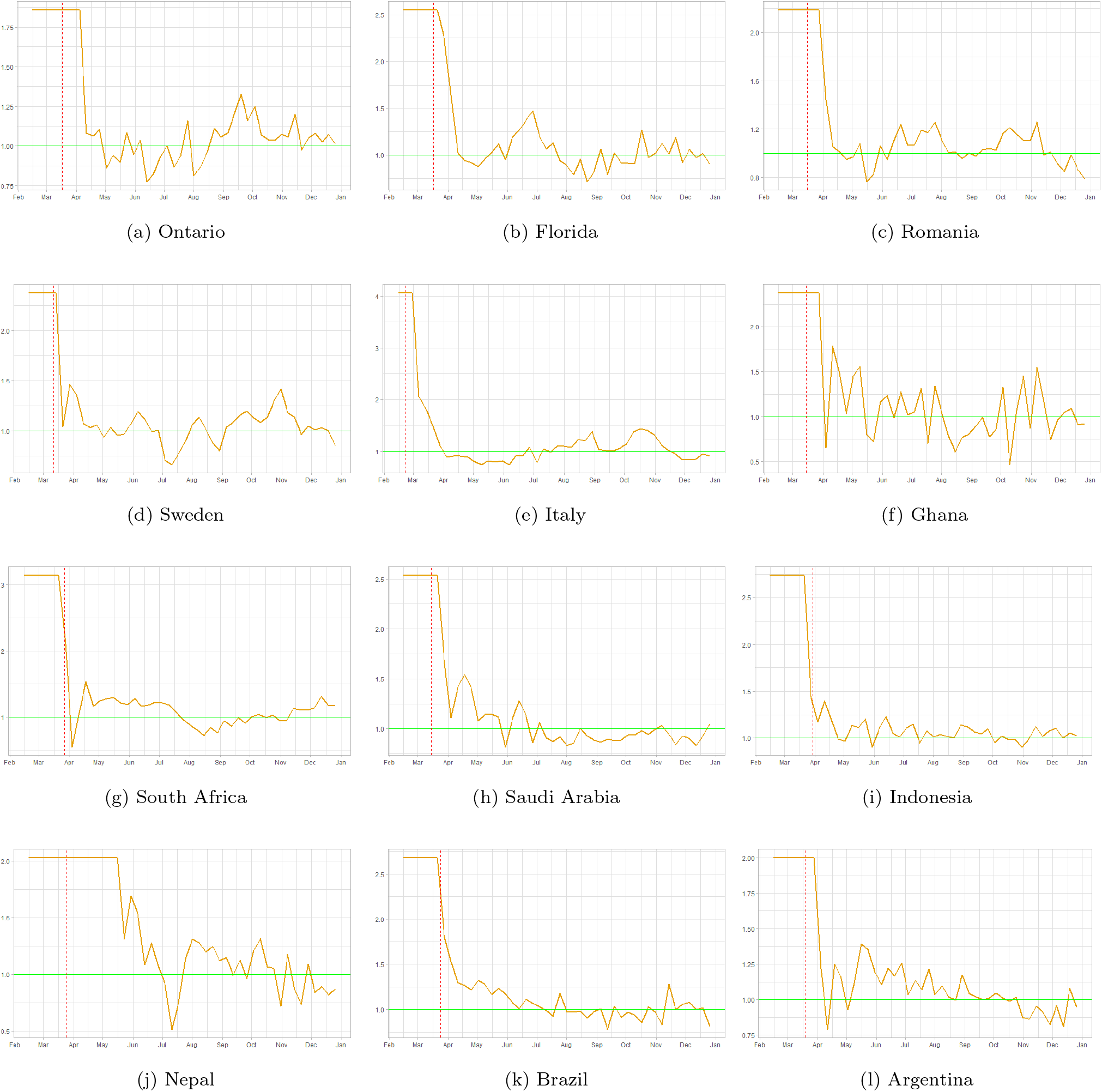
Weekly effective reproduction numbers based on incidence data. The yellow curve represents the outcome of equation (10), while the vertical pink dash line represents the start of nationwide social distancing orders in each region under consideration.

The structure of the paper is as follows: Section 2 presents the setup and methodology employed in the analyses of each of the twelve regions. Section 3 presents the results and discussion on quantification of the reduction in the effective reproduction numbers *R*_*eff*−*mobile*_ and *R*_*eff*−*mobilemask*_, as well as the estimated effective reproduction numbers based on incidence data *R*_*eff*−*data*_. The paper closes with a discussion of the results and a few directions for future work.

## 2 Methods and model setup

We chose 10 countries around the world, one U.S. state (Florida) and one province of Canada (Ontario), based on a diversity of characteristics: population density, median age, urbanization of population and gross domestic product (GDP) in 2020 (theses various characteristics are presented in Table 1 below:

**Table 1:** Countries under consideration.

|  | Regions | Population Den-<br>sity | GDP | Median Age | Urbanization |
| --- | --- | --- | --- | --- | --- |
| 1 | Ontario | 37.9/sq mi | 710 | 40.4 | 86.2 |
| 2 | Florida | 384.3/sq mi | 1095.9 | 42.5 | 91.2 |
| 3 | Romania | 218.6/sq mi | 248.6 | 42.5 | 56.4 |
| 4 | Sweden | 64.7/sq mi | 529.1 | 41.1 | 88 |
| 5 | Italy | 521.4/sq mi | 1848.2 | 46.5 | 71 |
| 6 | Ghana | 262.9/sq mi | 67.3 | 21.4 | 57.3 |
| 7 | South Africa | 109.8/sq mi | 282.6 | 28 | 67.4 |
| 8 | Saudi Arabia | 38.8/sq mi | 680.9 | 30.8 | 84.3 |
| 9 | Indonesia | 357.4/sq mi | 1089 | 31.1 | 56.6 |
| 10 | Nepal | 466.2/sq mi | 32.2 | 25.3 | 20.6 |
| 11 | Brazil | 64.7/sq mi | 1363.8 | 33.2 | 87.1 |
| 12 | Argentina | 37.3/sq mi | 382.8 | 32.4 | 92.1 |

The data are collected from [IndexMundi](https://www.indexmundi.com/factbook/countries). In each of these regions we are interested to model the pandemic evolution during the year 2020 via a SEIRL model described below.

Susceptible individuals (denoted by *S* and considered to be the entire population initially) exposed to the virus enter the exposed (*E*) compartment for an average of 1/*σ* days before they become contagious, at which point they move into the *I* (infected) compartment. In general, there is an *a* proportion of infected individuals who will not develop symptoms (so-called asymptomatic, denoted by *I*^*A*^), while the remaining 1 − *a* percentage make up the infected symptomatic individuals, denoted by *I*^*S*^. Thus here :

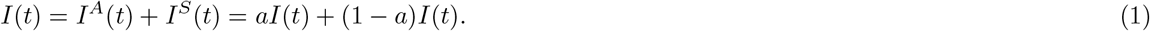

An *E* proportion of *I*^*S*^(*t*) will self-isolate into the *L* (isolated) compartment. They do so with a delay of 1/*κ* days, accounting for a test result wait time and/or individuals who may disregard minor symptoms initially. After 1/*γ* days, individuals recover from (or succumb to) their infection and move into the *R* (recovered) compartment. A diagram of the process we model is as follows 1:

Using the expressions in (1) and model diagram (1), we obtain the following equations:

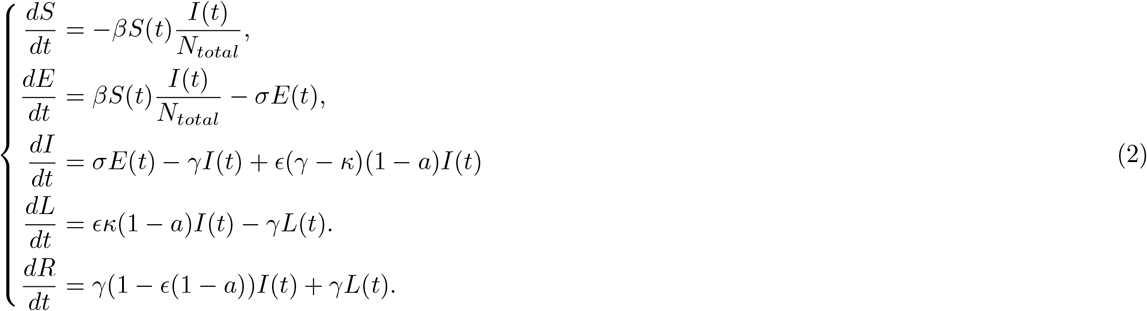

Our model parameters have been taken from literature (as can be seen in Table 2), with the exception of the rate of isolation *E*, which we assume to be equal to 95%.

**Table 2:** Parameter values for our model 2.

| Symbol | Definition | Initial Value | Reference |
| --- | --- | --- | --- |
| $N_{total}$ | Population size | Table 4 | |
| $\sigma$ | Rate at which exposed become infectious (days<sup>-1</sup>) | 1/2.5 | [36] |
| $a$ | Proportion of permanently asymptomatic cases | 0.5 | [37],[38],[39] |
| $\epsilon$ | Proportion of compliance with isolation | 0.95 | assumed |
| $\kappa$ | Isolation delay | 1 | assumed |
| $\gamma$ | Recovery/removal rate | 1/7 | [40] |

### 2.1 Time series of effective reproduction numbers using a near-disease-free equilibrium estimate and incidence data

The Jacobian analysis near the disease-free equilibrium (DFE) which consists of *S*(0) = *N* and *I*(0) = 0) for the system (2) and the next generation matrix method ([34]) for computing the initial *R*_0_ are given in Appendix. We have employed a similar type of approach on our papers [41, 42], where the compartmental models were slightly different. We obtain its closed form expression:

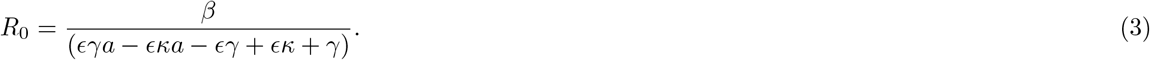

We can also estimate *R*_0_ as a function of the growth factor near the DFE in each region using (9) (see more details in Appendix):

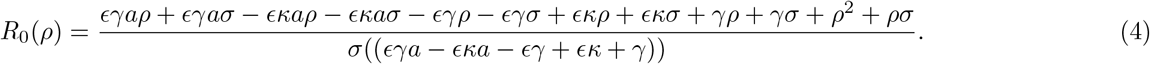

To estimate the exponential growth factor for each region as a time series, we rely on weekly case incidence data (which we denote by *inc*(*t*)) for each region. We assume that *inc*(*t*) is given by an exponential curve of the type:

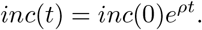

In this case, we can compute a time-series of the exponential growth factor:

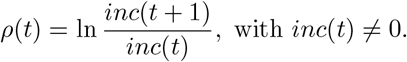

Using this time series of *ρ*(*t*) in equation (10) we obtain a time series for the effective reproduction number:

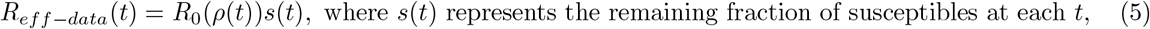

i.e. the difference between the entire population and the current cumulative number of infected individuals:

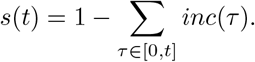

Figure 2 represents the weekly changes in the effective reproduction numbers of incidence data, *R*_*eff*−*data*_ that is explained in section (2.1), throughout 2020 for each geographical location under study. The vertical lines in each panel represent the dates in which local governments have introduced nationwide measures: in most countries lockdown measures took effect, while in Sweden and Indonesia partial lockdown measures were in place. In Sweden, nationwide lockdown was considered to be a violation of people’s freedom of movement.

The government strategy was based on individual responsibility. Measures included were: border closures, recommendations about social distancing, and traveling. Local reports show a 50% drop in public transport usage in April for the Swedish counties [43],[44]. In Stockholm, a 30% drop in the number of cars [45], and 70% fewer pedestrians [46] was reported in April 2020. Later in November, the Swedish government imposed mandatory restrictions as well (e.g. all gatherings of more than eight people were banned).Italy had one of the earliest implementations of lockdown, as early as February 23, 2020. We showcase all dates for all locations in Table 3.

**Table 3:** Nonpharmaceutical Interventions (NPIs) measures

|  | Regions | Hard/partial Lockdown order | Mask Mandate |
| --- | --- | --- | --- |
| 1 | Ontario | 17 March | 17 July |
| 2 | Florida | 17 March | 25 June |
| 3 | Romania | 16 March | 1 August |
| 4 | Sweden* | 11 March | 7 December |
| 5 | Italy | 22 February | 7 October |
| 6 | Ghana | 15 March | 15 June |
| 7 | South Africa | 27 March | 1 May |
| 8 | Saudi Arabia | 15 March | 22 May |
| 9 | Indonesia | 30 March | 5 April |
| 10 | Nepal | 24 March | 31 July |
| 11 | Brazil | 24 March | 2 July |
| 12 | Argentina | 19 March | 29 August |

### 2.2 Data sources

In the remainder of the paper we use various sources of publicly-available data. First we use Google data mobility [47] for each region of interest. Then we use Johns Hopkins data for case incidence: incidence [35] and we use the Prem et al. [21] paper and their contact data projections (where projections for provinces or states in Canada and the US were obtained by using the projections weighted with population data for such provinces or states. Lockdown measures start dates have been taken from the ascent of the Oxford stringency index at Financial Times.

Population data were obtained from Stats Canada [48], US Census data [49], whereas for other countries we used the online repository: World age distribution [50]. For obtaining data on mask wearing compliance we used Mask Compliance [51] and European-countries [52].

### 2.3 Time-series of effective reproduction numbers accounting for mobility data and projected contact rates

We first devise a mechanism to mix the daily average contact rates in each region’s population with the behavioural activity in that population. In Prem et. al. [21] the authors compute projected daily average contact rates for 157 countries. Specifically, they estimate the average contacts for categories of activities during a typical day, such as: home, work and other locations. They present their results for a population stratified by age, and divided into 16 age subgroups (See Appendix for the details). We amalgamated the average contact rate in home, work, and other locations, then we computed the weighted average of a given projected contact matrix in a region with the corresponding proportions of 5-year age population groups in 2020 to determine one single average contact rate. We consider this last value as the average baseline (pre-lockdown) contact rate in that region (for instance, for Ontario it was computed to be: *contact*_*av*_ *≈* 11.04). All regions’ contacts are reported below in Table 4.

**Table 4:** The basic reproduction number, *R*_0_, at the beginning of the COVID-19 pandemic and contact average rate, *contact*_*av*_, before the COVID-19 outbreak for each geographical location under study. The table also contains total population numbers and an estimated level of mask compliance in each population - data sources highlighted in Section 2.2.

| | Region | Population<br>$N_{total}$ | $contact_{av}$ | $R_0$ | $\rho$ | # of<br>weeks ($\rho$) |
| --- | --- | --- | --- | --- | --- | --- |
| 1 | Ontario | 14,734,014 | 11.04531438 | 1.856251 | 1.239646 | 5 |
| 2 | Florida | 21,477,737 | 10.50481103 | 2.547635 | 2.044037 | 2 |
| 3 | Romania | 19,237,682 | 10.53032815 | 2.183026 | 1.63537 | 3 |
| 4 | Sweden | 10,099,270 | 11.09614737 | 2.372179 | 1.851232 | 4 |
| 5 | Italy | 60,461,828 | 12.38928992 | 4.054303 | 3.485834 | 2 |
| 6 | Ghana | 31,072,945 | 16.04056691 | 2.373496 | 1.852704 | 2 |
| 7 | South Africa | 59,308,690 | 13.21954859 | 3.143281 | 2.654134 | 2 |
| 8 | Saudi Arabia | 34,813,867 | 13.26255761 | 2.533263 | 2.028494 | 2 |
| 9 | Indonesia | 273,523,621 | 12.54859287 | 2.733693 | 2.241501 | 2 |
| 10 | Nepal | 29,136,808 | 16.09865556 | 2.025042 | 1.447956 | 2 |
| 11 | Brazil | 212,559,409 | 12.72484009 | 2.680081 | 2.185299 | 2 |
| 12 | Argentina | 45,195,777 | 12.13054343 | 1.998173 | 1.415379 | 3 |

Google reports contain changes in movement over time, compared to baseline (pre-lockdown) activity in six categories: retail/recreation, groceries/pharmacies, parks, transit stations, workplaces, and domiciles (COVID- 19 Community Mobility Reports) [47]. We have used the Google index data in other works, see [20, 53]. To find the mobility-influenced, time-dependent contact rates post-lockdown, we considered the average contacts rate for the home, work, and other location categories (comprising retail/recreation and groceries/pharmacies) from Prem et al. [21] for each region. Next, we used these category rates to modify the same categories of mobility data as follows:

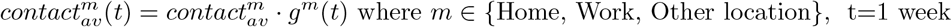

and where *g*^*m*^(*t*) is the percentage increase or decrease in the category *m* of mobility as compared to Google’s baseline values per category. Finally, we amalgamated the Google mobility-influenced contact rates of these categories to compute the weekly mobility-influenced contact rate by considering an average number of weekly hours *hr*_*m*_ in each category:

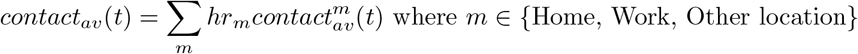

Let us now refine our view of the effective reproduction number *R*_*eff*_ = *R*_0_*s*(*t*) from the perspective of the changes in contact rates due to mobility reduction. From the last section we know that 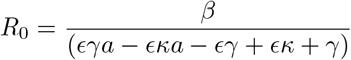, however now we have

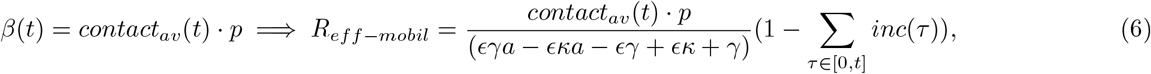

where *p* is the probability of transmission per meaningful contact. To estimate *p*, we need to have a handle on the values of *R*_0_ from incidence data as well. To estimate *R*_0_ from incidence data we used equation (10). The growth factor *ρ* is computed from the initial phase of (close to) exponential growth in the neighbourhood of the DFE, which corresponds to a phase of linear growth in *log*(*inc*), with slope *ρ*. We identify the initial, fastest phase of nearly unchecked growth in any given region with the help of a piecewise linear fit to the log of the incidence. We utilize the `R` function `dpseg()`, which is a part of the `dpseg` package, https://cran.r-project.org/web/packages/dpseg/index.html. This function uses a dynamic programming algorithm to generate an optimal piecewise linear fit to a time series, which balances goodness of fit against an (adjustable) penalty for each additional segment. We then identify the earliest segment with the steepest positive slope (largest *ρ*) as corresponding to the initial near-unchecked exponential growth phase. We report the values we obtain in Table 2.^[2]^

Table 4 summarizes the basic reproduction number *R*_0_ at the beginning of the COVID-19 outbreak and the average contact rate (before pandemic) and mask compliance level (*compliance*_*m*_). The basic reproduction number *R*_0_ has maximum values in Italy and South Africa with 4.05 and 3.14, and minimum values in Ontario and Argentina with 1.85 and 1.99. We retrieve different mask compliance levels for each region from different sources (see Section 2.2).

## 3 Results and Discussion

### 3.1 Quantifying the effective reproduction number using incidence data

To quantify the overall relative status of the pandemic at the various locations, we show in Figure 3 the evolution of *R*_*eff*−*data*_ in each region using a heatmap plot and depicting values of *R*_*eff*−*data*_(*t*) ∈ [0, 4].

**Figure 3:**
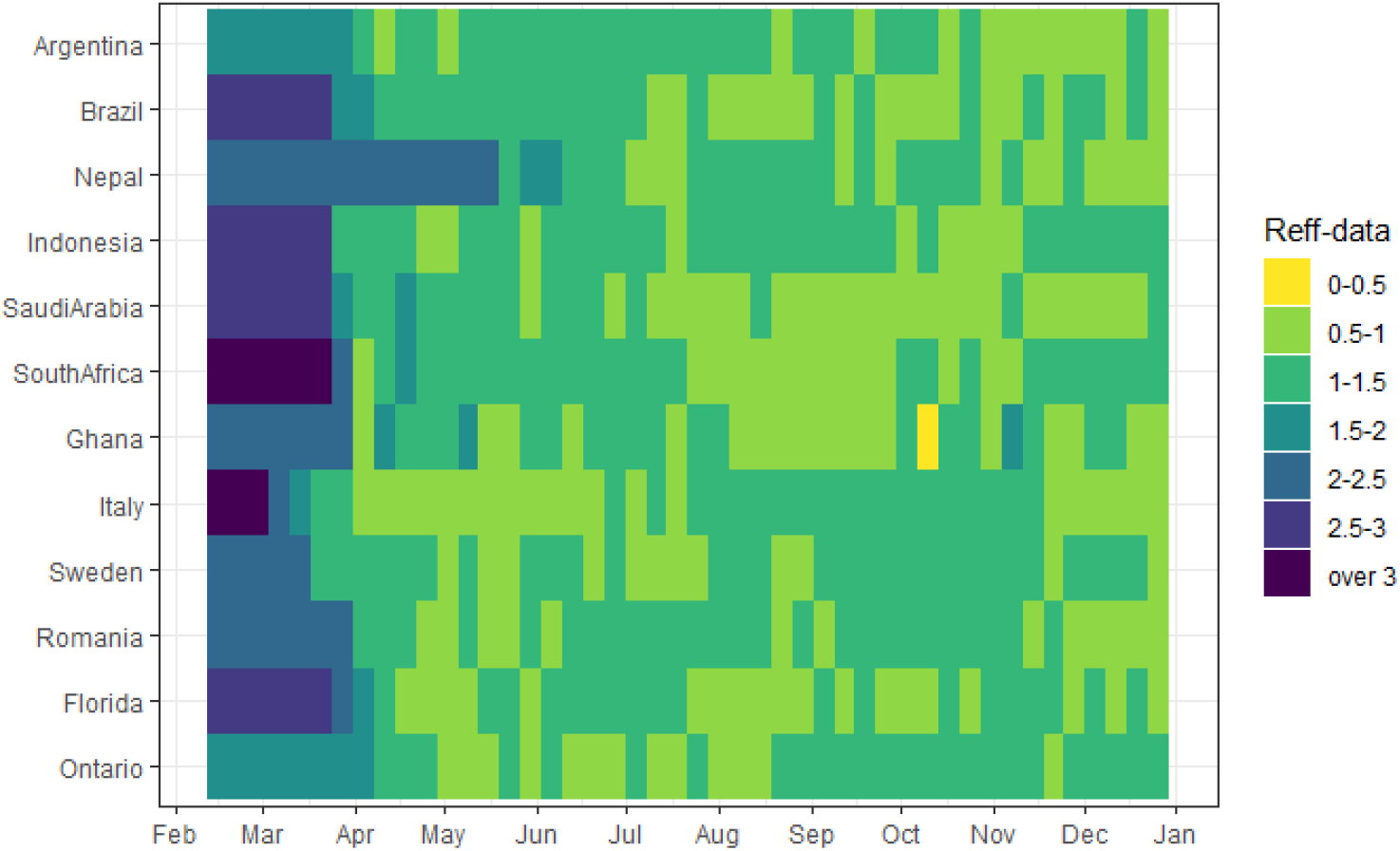
The effective reproduction numbers from incidence data plotted per range of values (*R*_*eff*_ *∈ {*[0, 0.5], [0.5 − 1], …, [2.5 − 3]*}*) from February 15 to December 31, 2020. Lightest color patches signify biggest reductions in values of *R*_*eff*_. In the second panel we see the cumulative incidence for the same regions

The color gradient signifies lowest values of the effective reproduction numbers in lightest color zones, and highest values in darkest zones. Figure 3 showcases a comparison of the *R*_*eff*−*data*_ between locations. We can immediately see the effect of the reduction in *R*_*eff*−*data*_, across the board, in all countries between March and April, and the rest of the year. It is clear the drop achieved by initial lockdowns and mobility reduction allowed all countries to drop their *R*_*eff*−*data*_ to around 1, and interestingly most remained around this value for the rest of 2020. We include here (Table 5) the average *R*_*eff*−*data*_ at each location *after* the initial drop.

**Table 5:**
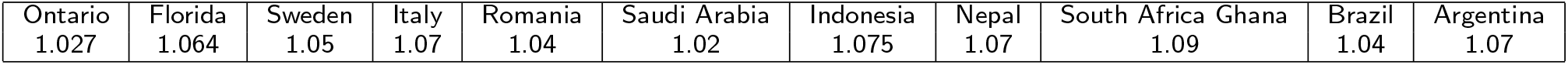
Average *R*_*eff*−*data*_ at each location after the initial drop

Qualitatively, we can understand why *R*_*eff*−*data*_ has generally fluctuated in the vicinity of 1: On the one hand, rapid growth of incidence causes alarm at both the policymaker and individual level, and typically leads to increased NPI directives as well as compliance. On the other hand, because these measures are hard to maintain and economically taxing, NPIs are generally relaxed not long after incidence starts to drop. In other words, it is the human behavioural element that we do not model here, but that we glimpse.

### 3.2 Quantifying the effective reproduction numbers accounting for mobility data

We observe that the initial sharp reduction in contact rate due to mobility happened in the middle of March in most regions under study, when initial lockdown was in effect, except in Sweden. The contact rate gradually increased from early May when partial reopening was implemented by governments. The results of these measures are noticeable in the figure 4 and 7 upper right panels.

**Figure 4:**
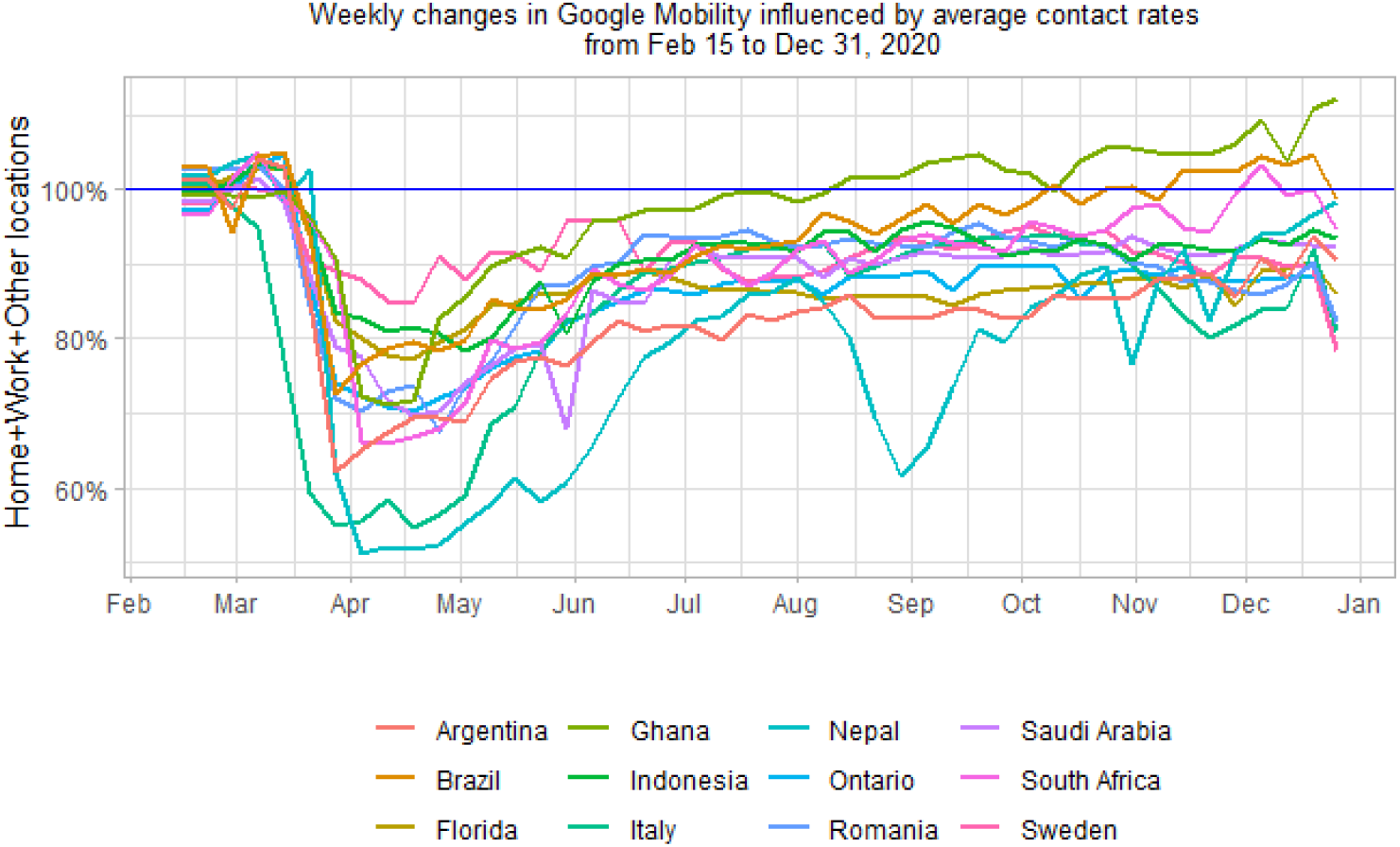
Weekly amalgamated Google Mobility Index in the Home, work and other activities respect to the baseline (pre-lockdown) influenced by average contact rates, from Feb 15 to Dec 31, 2020. Baseline rates were computed from Google mobility reports over a 6week interval of January-February 2020.

The effect of mobility restrictions throughout 2020 for each geographical location under study is presented in Figure 5 below. We plot the theoretically estimate *R*_*eff*−*mobile*_ numbers together with the *R*_*eff*−*data*_ effective numbers at each location. Overall, under Google mobility reductions using formula 6, the mobility reduction effect is not enough, on its own, to explain the values *R*_*eff*−*data*_. This is not surprising, as each location had a varying combination of NPI measures.

**Figure 5:**
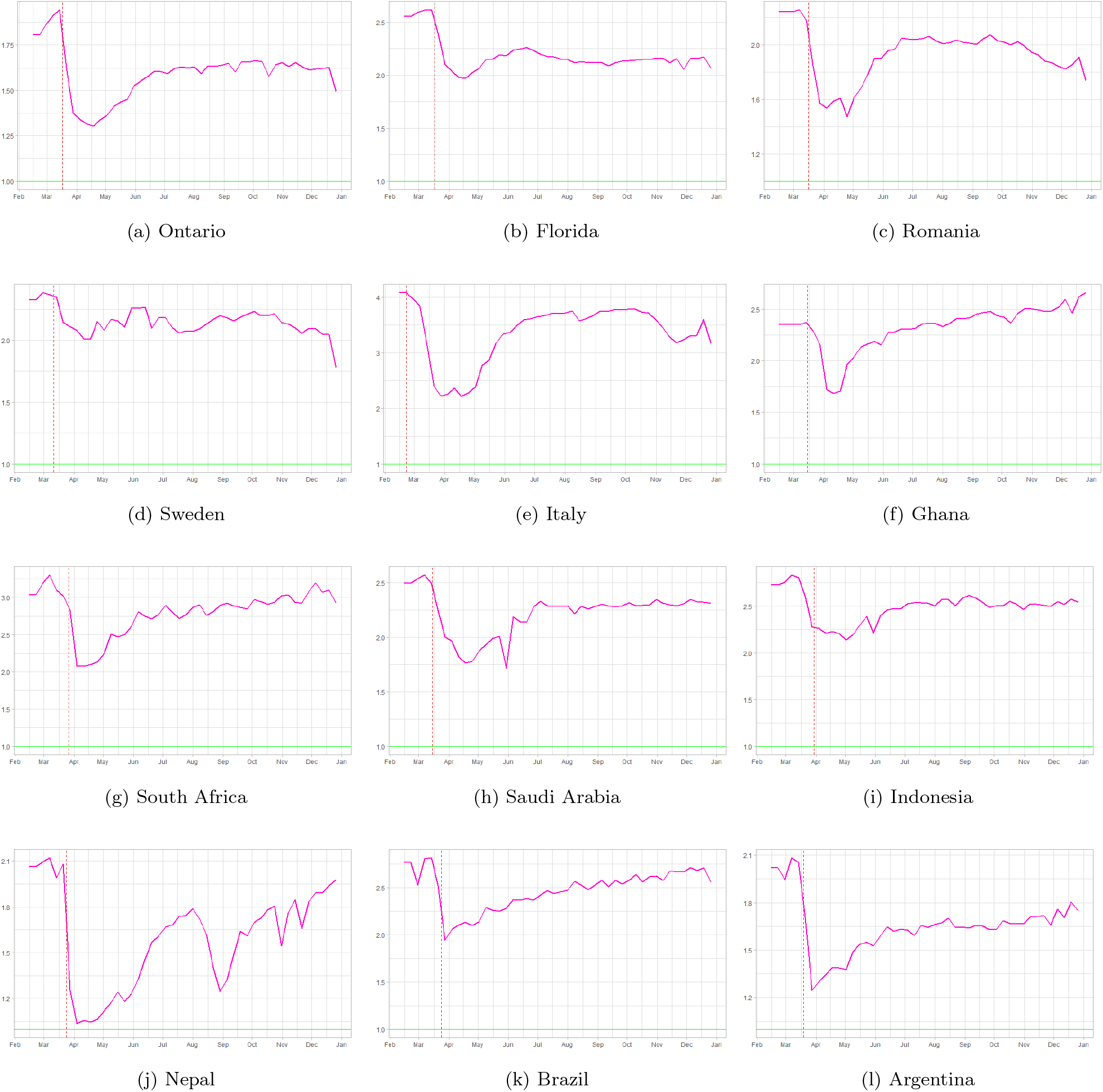
Effective reproduction numbers from the mobility data influenced by contact rate. In each panel, the red vertical dash line represents the lockdown measures date and the pink curve shows the weekly effective reproduction numbers from the mobility affected by average contact rate in that region.

**Figure 6:**
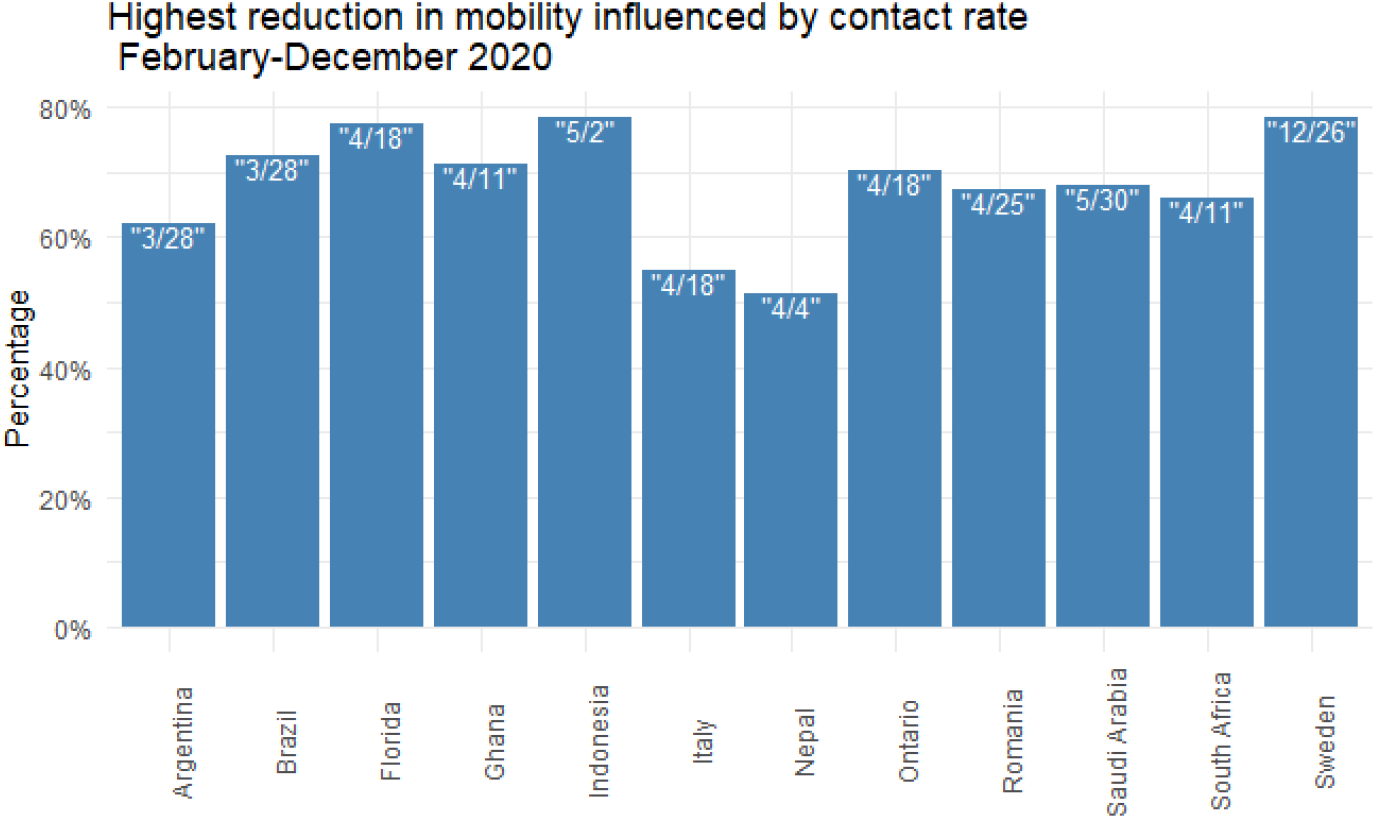
Reduction in mobility influenced by contact rate throughout 2020. The week that reduction happened is highlighted by “month/day” in each bar.

Figures 5 (and 6 in an ensemble view) reveal the reduction in mobility in each region. It seems that the mobility decreased 51% and 54% in Nepal and Italy with respect to the baseline in the first and second weeks of April 2020, respectively. The mobility fell about 20% percent in Indonesia and Sweden. Large-scale social restrictions (sometimes called partial lockdown) were introduced by the Indonesian government in place of nationwide lockdown at the end of March 2020 while the Swedish authorities imposed some restrictions on gatherings in late November.

### 3.3 Human behaviour and its impact on disease transmission

#### 3.3.1 The effects of mask mandates and mask compliance on further reduction of the effective reproduction numbers

We will be looking at two main tools that populations can use to control the pandemic: *social distancing* and *mask wearing*. In our framework here, each one of these directly influences the transmission rate *β*. Denoting by *mask*_*eff*_ the efficacy of an average mask at preventing transmission and by *compliance*_*m*_ the compliance with mask wearing let us imagine a meaningful contact between an infected and a susceptible individual: If an individual wears a mask and *mask*_*eff*_ = 0.3, then he/she has an increased protection against transmission. If the individual complies with mask wearing 50% of the time, then their protection due to mask wearing is, on average *mask*_*eff*_ · *compliance*_*m*_ = 0.15 which implies that the per-contact transmission probability *p* will decrease with mask wearing:

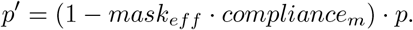

Recalling that the level of mask-wearing and social distancing both change over time, we estimate that

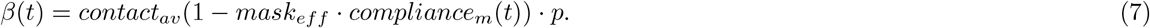

Let us consider only mask-wearing for the time being. In this paper we use a value of *mask*_*eff*_ = 50%, as averaged based on estimates in [54] (see a more detailed discussion in Appendix. and the values of mask compliance from IHME (details of data sources in Data sources section above.). Using (7) in the estimate (6) leads to:

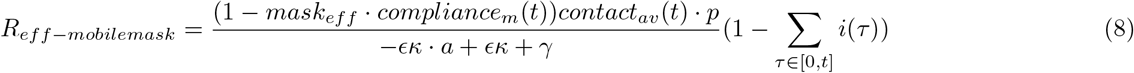

with data from Table 4.

Figure 7 shows the effective reproduction numbers as obtained from the incidence data (yellow curves) from Figure 2, together with our theoretical estimates from Figure 5 (solid pink curves) using mobility data, to which now we add a new theoretically estimated *R*_*eff*−*mobilemask*_ number reflecting mask wearing data and formula 8 above. The red and black dashed lines in each panel show the lockdown and mask-wearing mandate dates, respectively, in each region as publicly available.

**Figure 7:**
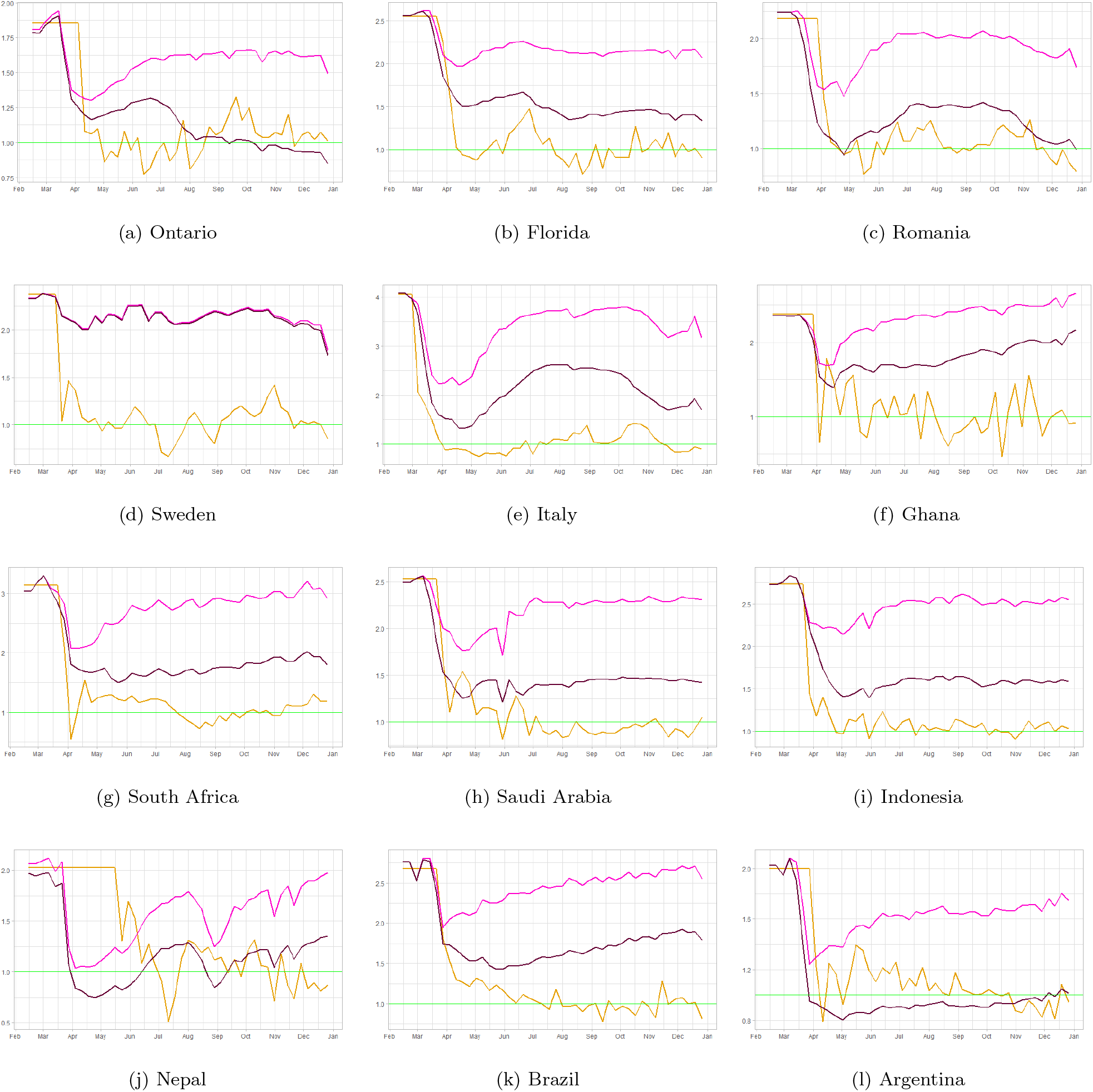
A comparison of the effective reproduction number as obtained from the incidence data (yellow curves), to our theoretical estimate from Section 3.1 (solid pink curves) using mobility data. Beyond the date of mask mandate enactment in each region, we show the theoretically-estimated incidence both with (solid pink) and without (dashed pink) the added effect of mask-wearing. The red and black dashed lines in each panel show the lockdown and mask-wearing mandate dates, respectively, in each region as publicly available.

We observe that the theoretically-estimated curve using both mobility reductions and mask wearing data (which we will now denote by *R*_*eff*−*mobilemask*_) is accounting for more of the reduction in transmission than the estimated *R*_*eff*−*mobile*_ curves of Figure 5. Evidently, in some regions (Ontario and Argentina) we observe what it looks like an over-reduction in the values of *R*_*eff*−*mobilemask*_ as compared to *R*_*eff*−*data*_, while in other countries, most notably Sweden, we notice essentially no contribution in further reduction from *R*_*eff*−*mobile*_ to *R*_*eff*−*mobilemask*_. In Sweden’s case, this is not surprising, as the mask wearing levels in the IHME data we used hover between 1% − 2% throughout 2020. Last but not least, this assumes a values of mask efficacy *mask*_*eff*_ = 50%. If this value is decreased, the *R*_*eff*−*mobilemask*_ curves will shift upwards, i.e. the reduction of 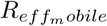 due to mask wearing will be smaller.

#### 3.3.2 Effects of other social distancing measures on further reduction of disease spread

As we highlighted in previous section, a host of other NPI’s have been employed, at different strengths, across the regions. While it is clear from our investigations thus far that the mobility reduction (as reflected in Google mobility data), along with mask wearing and mask compliance helped tremendously in the de-escalation of the *R*_*eff*_ curves, we wish to further analyze the data to extract more information on the strength of other NPI’s. Let us denote by *compliance*_*oNPI*_ the compliance level in the local population with *other NPI* measures (by other we mean other than mobility and mask wearing). With this in mind, an average individual in a local population is protected over the course of their contacts proportional to what fraction of the time they also comply with other NPI measures:

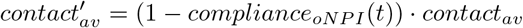

which then means that

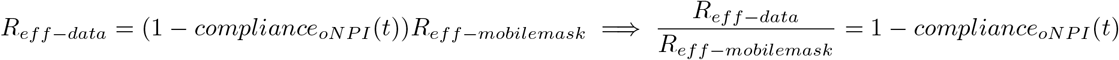

This means that to quantify the effects of other NPI measures per region, we further look at the ratio: 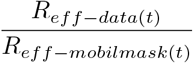. We present these plots in Figure 8 below. Clearly when the ratio is less than 1, then the reduction in the *R*_*eff*_ that we quantified from observed data is stronger than the reduction of *R*_*eff*_ based on mobility and mask wearing and viceversa. Specifically we obtain the following:

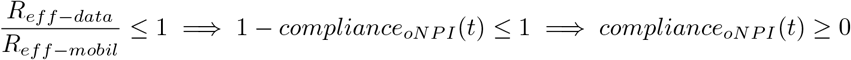

**Figure 8:**
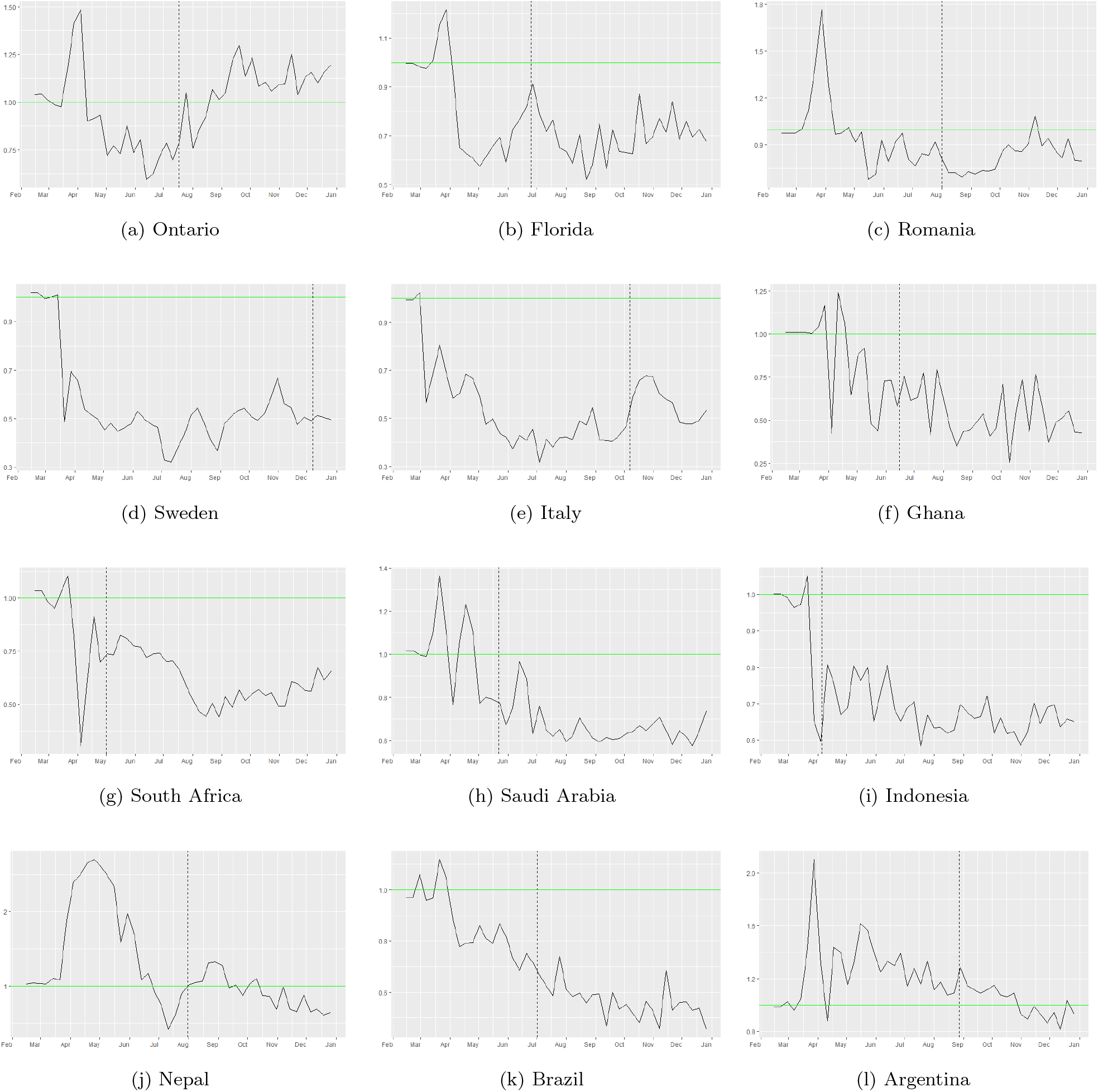
Effect of other NPI measures (e.g. 6 feet (2m) social distancing, hand washing, etc.). Mandatory masks were introduced at dates represented by the vertical dashed line in each panel.

In the other case, under our assumptions, we get

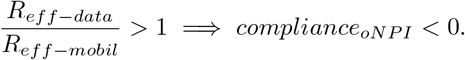

In cases where *compliance*_*oNPI*_ < 0 we interpret it to mean that while the local population has been very compliant with mask wearing (assumed to be at 50% efficacy in all regions), they may not have been as observant towards other measures.

One remark is worth to be made at this point: if one decreases the mask efficacy *mask*_*eff*_ to an average of 40%, respectively even lower to 30% (as in [54]), then the *R*_*eff*−*mobilemask*_ curves of Figure 5 would scale equivalently upward by a constant, thus implying that the mask wearing effect is less effective and therefore that the ratio 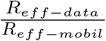 would be less than 1 (i.e., *compliance*_*oNPI*_ (*t*) > 0) for most (respectively all) regions.

Results in Sweden in particular stand out (see Figure 9). They indicate that other NPI measures were extremely effective in reducing the transmission rate of disease, in the absence of mandated lockdown periods and nearly 0 mask wearing. In the case of Sweden for instance (upper panel of Figure 9), *compliance*_*oNPI*_ (*t*) is large starting in April 2020. Here *compliance*_*oNPI*_ (*t*) > 0 at and over 50% in Sweden. In case of Indonesia, we estimate that *compliance*_*oNPI*_ (*t*) at and over 30-40% in Indonesia. Both these ranges assume an average of *mask*_*eff*_ = 50% over the time period May-December 2020.

**Figure 9:**
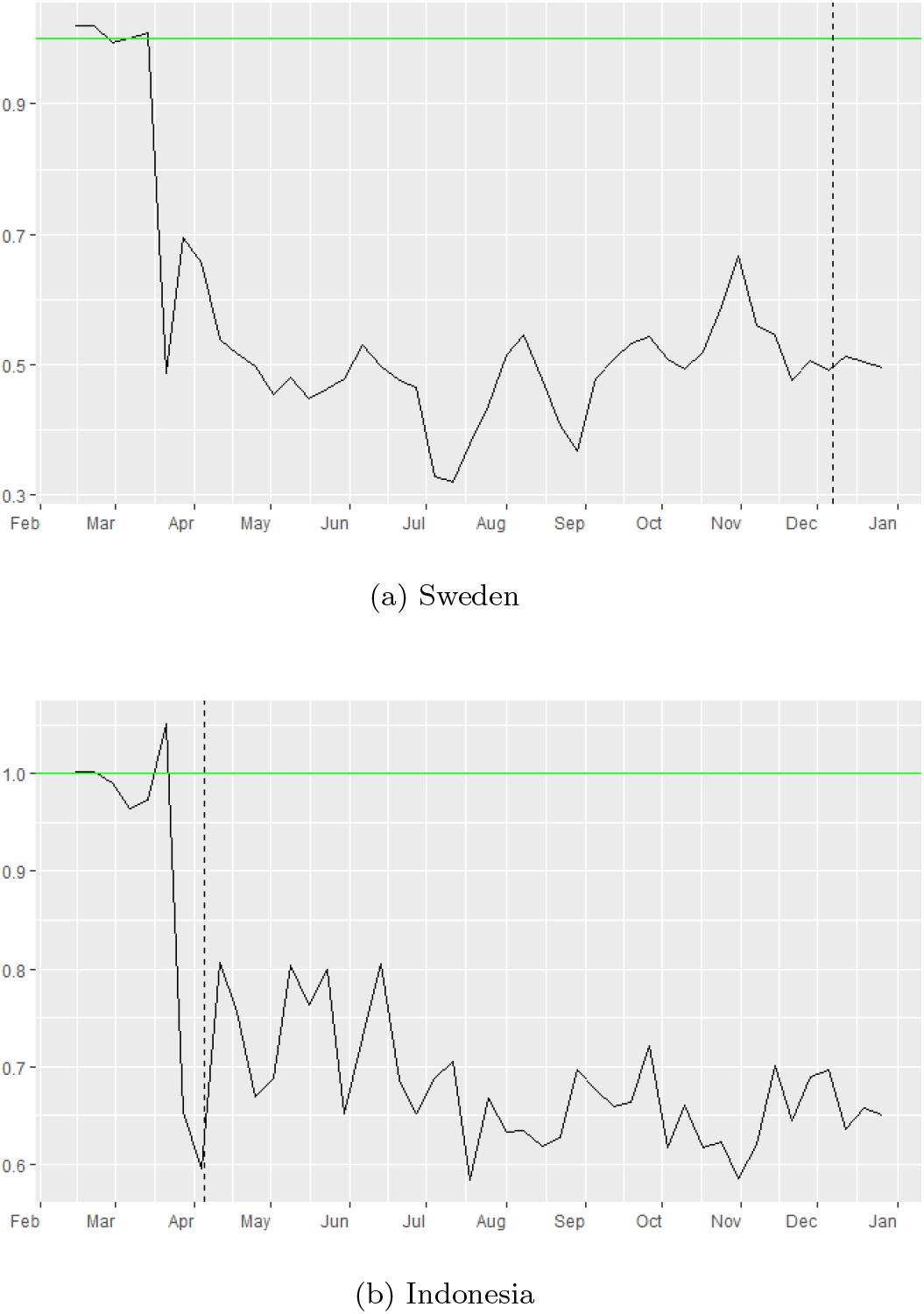
Effect of other NPI measures in Sweden and Indonesia. Mandatory masks were never introduced in Sweden until November 2020, but they are present earlier in Indonesia. Nationwide lockdown was never used in these countries in 2020.

At the other end of the spectrum, Ontario (upper left corner panel in Figure 8) seems to display a ratio of 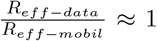 after the mask mandate date (vertical black dash line). Moreover, we have periods of time where the ration is larger than 1 somewhat, so then *compliance*_*oNPI*_ might have been negative (in the case where *mask*_*eff*_ = 50%). This seems to indicate that, under our assumptions, the mobility reductions captured via Google indexes and the mask compliance levels have essentially captured the full picture of the pandemic evolution in this region. Similar trends can be seen in Argentina and Nepal (see Figure 8 and Figure 10).

**Figure 10:**
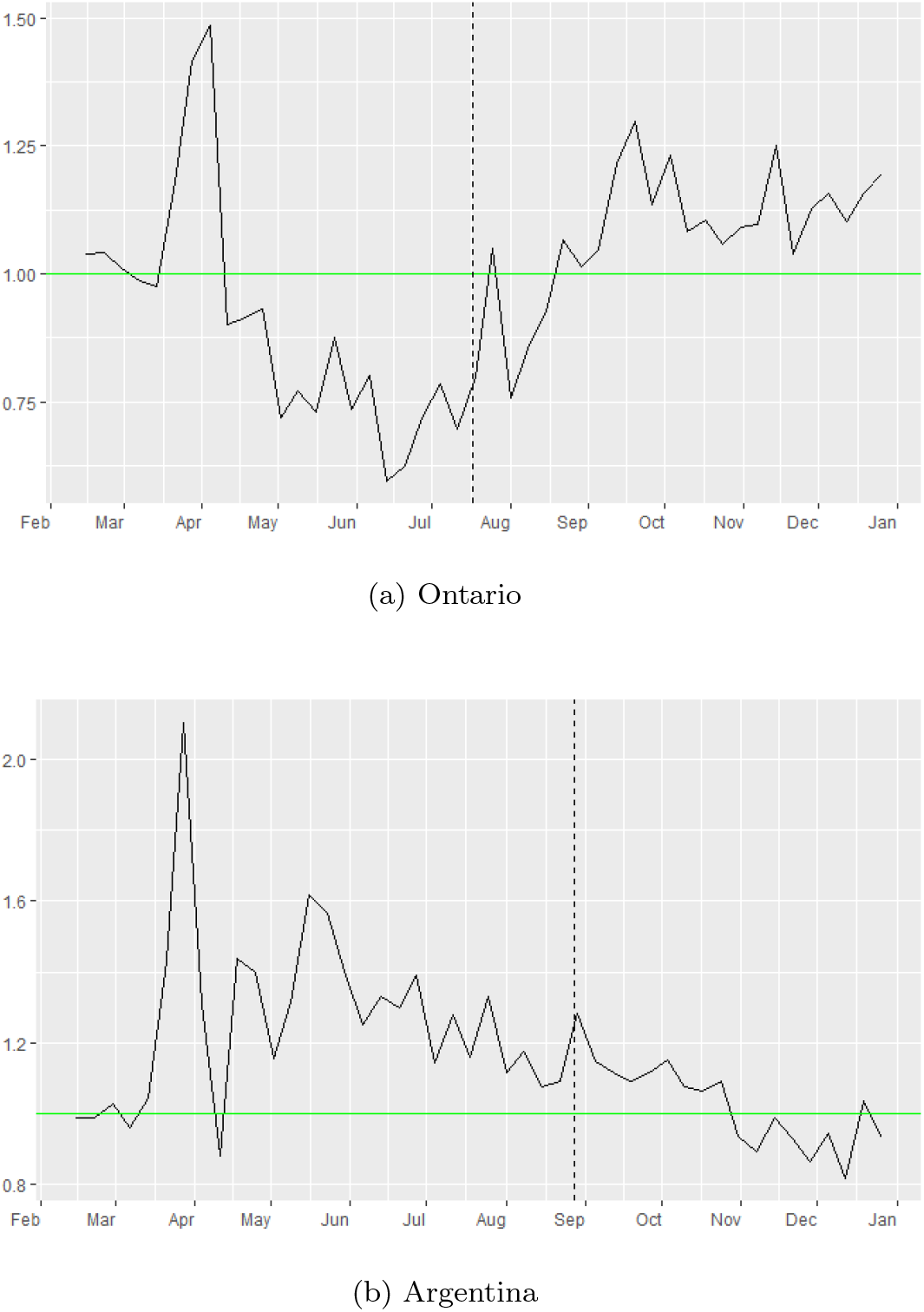
Effect of other NPI measures in Ontario and Argentina. Mandatory masks were introduced at dates represented by the vertical dashed line in each panel.

#### 3.3.3 Further investigations into mask efficacy and its impact on transmission

In the sections above, we have considered a fixed value of *mask*_*eff*_ = 50% across all regions, and we have commented on how a decrease of this value may affect the reductions of *R*_*eff*−*mobile*_ values. From formula 6 we have

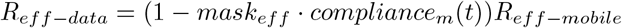

for each region. Here, we perform a maximum-likelihood estimate of *mask*_*eff*_ for each region, such that the fit of *R*_*eff*−*mobile*_ to *R*_*eff*−*data*_ is optimized. In other words, if we considered NPIs to consist only of mask-wearing, what mask efficacy would best reproduce the observed time series of effective reproduction number in a given region? We present our results in a new plot with a small Table 6:

**Table 6:** Maximum likelihood best fit values for *mask*_*eff*_ per country

|  |  |  |  |  |  |  |  |  |  |  |  |
| --- | --- | --- | --- | --- | --- | --- | --- | --- | --- | --- | --- |
| Ontario | Florida | Sweden | Italy | Romania | Saudi Arabia | Indonesia | Nepal | South Africa | Ghana | Brazil | Argentina |
| 39% | 75% | 100% | 83% | 52% | 80% | 82% | 45% | 85% | 98% | 88% | 44% |

The graphs of the new *R*_*eff*−*mobilemask*_ time series are presented in Figure 11 (dark pink curves). Here we see again that Sweden stands out simply because mask wearing was not a policy the population had adopted in 2020; even at a 100% efficacy, an adoption of 1-2% has negligible effect. Thus other NPI factors must have been in play.

**Figure 11:**
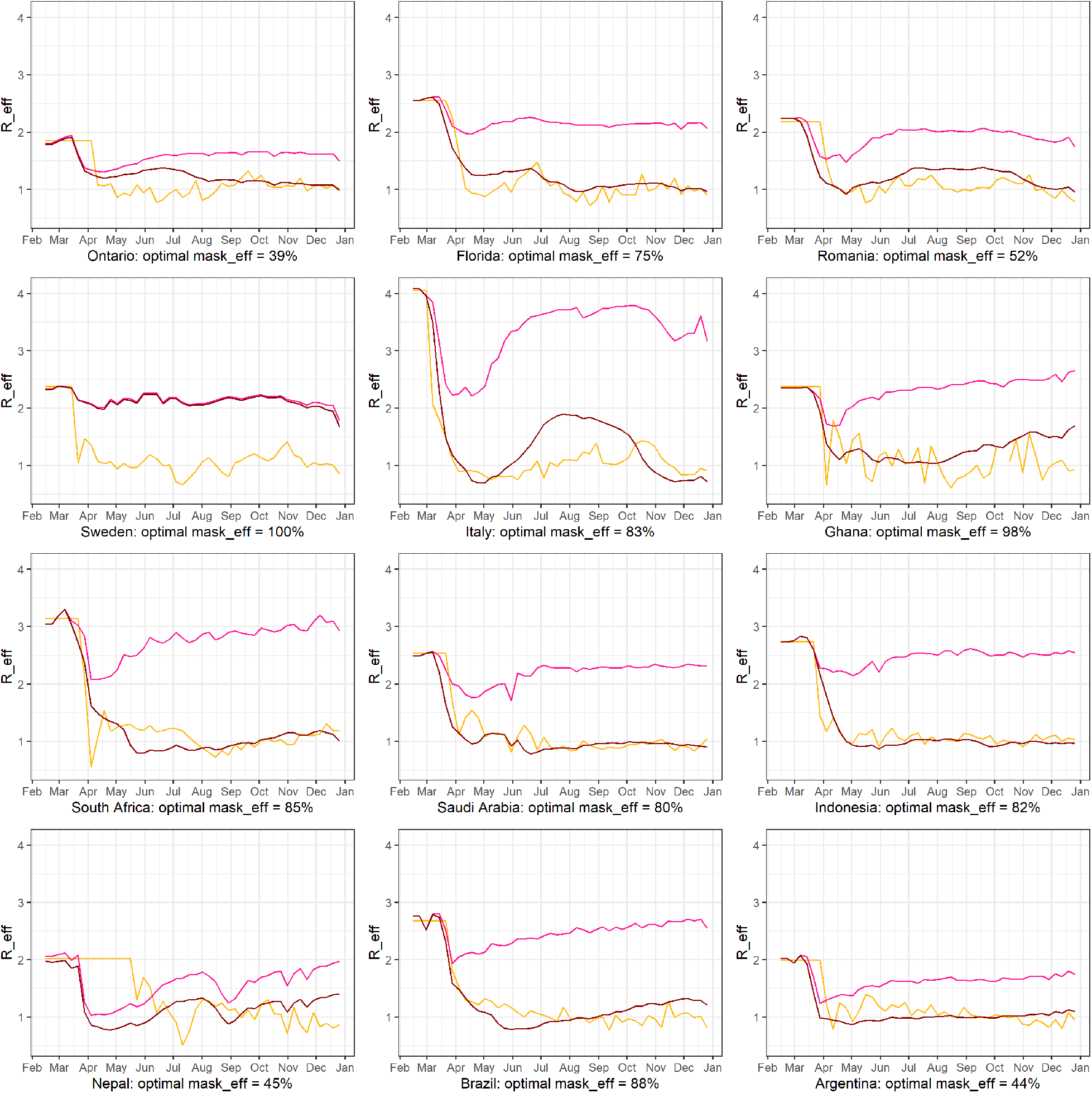
Maximum-likelihood estimate of *mask*_*eff*_ for each region so that the fit of *R*_*eff*−*mobile*_ to *R*_*eff*−*data*_ is optimized. Curves in dark pink color are the new *R*_*eff*−*mobilemask*_ estimates, where for each country we use the deduced value of *mask*_*eff*_ listed.

Among all the other countries, we see that high levels of mask wearing (for instance in Ontario, and similarly Romania and Argentina) correlates to realistic mask efficacy values (in [54] a realistic expected protection from the mostly cloth-type masks available widely in 2020 is between 30% to 50%) In regions such as Florida, Italy, Indonesia, South Africa, Ghana and Brazil the lower mask wearing levels would have needed to be compensated by higher mask efficacy levels. Since such levels of mask efficacy were not possible in 2020 for an average individual in any of these countries, then other NPI factors had to have been in play.

This analysis furthers the importance of our analysis in the last subsection (Subsection 3.3.2) where we manage to quantify the effect of other NPI factors (*oNPI*) in reductions of transmission in each of the 12 regions.

## 4 Conclusion

This paper shows that estimated epidemiological parameters of an underlying SEIR(L) model can be used to compute a time series of effective reproduction numbers accounting for mobility and mask wearing (explicitly using available free data), which are then used to highlight differing pandemic trajectories in very different parts of the world. Our sample consisted of 12 regions (10 countries, 1 Canadian province and 1 US state) chosen based on diversity of population density, median age, urbanization of the population, projected average contact rates, and gross domestic product (GDP) as in Table 1. We used a SEIR(L) epidemiological model (with parameter values from existing literature) and we used it to compute the near disease-free mathematical expressions of the effective reproduction numbers in terms of initial exponential growth of infection. While we made some specific choices on the structure of the compartments and the flow rates between them, we note that the mathematical methodology we applied is universally applicable to other forms of SIR and SEIR models, and not just to ours. This makes our results applicable to many other variants of compartments models for infectious disease transmission, not just for SARS-Cov-2 (for instance the next concern in the wake of the pandemic are the next flu seasons and the possible mitigation measures in case high;y infectious flu variants will make an appearance).

We were able to highlight quantitative relationships between the inferred weekly effective reproduction numbers and the estimated weekly effective numbers based on mobility reduction (as captured by Google mobility index) and mask-wearing (as captured from existing data).

Figures 2 and 3 show that there is a sharp drop in *R*_*eff*−*data*_ at the initial lockdown, then most countries have maintained their effective reproduction numbers around 1 by a combination of reduced mobility, mask-wearing and some additional other NPI’s not. Figure 7 shows a mobility-induced decrease of *R*_*eff*−*mobil*_ with a further decrease when we add mask wearing levels in each region.

Further, our modeling analyses provide direct illustrations of the effectiveness of other NPI’s in controlling transmission in each region. Figures 7 and 8 show that effective reproduction numbers in all regions have been helped by population adherence and practice of NPI’s over and above reductions in mobility and mask protection. This is also obvious by comparing the values of the effective reproductions numbers inferred from data versus (in Figure 7 most regions maintain their values in the neighborhood of 1), while mobility reduction alone (as illustrated from Google mobility reports and in Figure 4) has steadily decrease beyond May 2020 in all locations.

There are several assumptions underlying our study. Clearly, the mobility reduction as reflected in Google mobility reports is used here as representative across each of the populations, however that may not be quite accurate and it depends on the percentage of cellphone usage in a region and whether or not that percentage can be considered representative of an average individual. At the same time, the pre-pandemic projected contact rates from [21] are themselves estimates, thus subject to further change or calibration, given the wealth of data from last year studies.

Nevertheless, the overall ideas we followed are fairly straightforward and the quantification of the control on the pandemic via time series of effective reproduction numbers can be done as shown by using parameter values and data, without the need to model and fit SEIR model curves. Even if some of the assumptions/values in our analysis change, the methodology we propose is flexible, novel and easy to follow and implementing any new or updated piece of data available is straightforward. As future directions of research we are interested in using differing data sources for contact rates in some regions and differing estimates for mobility reductions to replace the Google mobility reports. We can also us differing *S*(*E*)*IR* models and more data on socio-economic and demographic factors that may lead to further insights into accounting for differences in pandemic evolution in diverse countries around the world.

## Data Availability

The data used and/or analyzed during the current study are derived from public domain resources. The datasets are available in the [COVID-19 Community Mobility Reports] repository, {https://www.google.com/covid19/mobility/}, the [COVID-19 Data Repository by the Center for Systems Science and Engineering (CSSE) at Johns Hopkins University],{https://github.com/CSSEGISandData/COVID-19}. We used the [Prem et al. contact data], {https://journals.plos.org/ploscompbiol/article?id=10.1371/journal.pcbi.1005697#sec020}. Population data were obtained from Stats Canada {https://www.statcan.gc.ca/eng/start}, US Census data {https://data.census.gov/cedsci/table?q=Florida&tid=ACSST1Y2019.S0101&hidePreview=false}, and {https://population.un.org/wpp/Download/Standard/Population/} for other countries. The data related to mask wearing compliance are available on {covid19.healthdata.org} and {https://www.statista.com/statistics/1114375/wearing-a-face-mask-outside-in-european-countries/}.

https://www.google.com/covid19/mobility/

https://github.com/CSSEGISandData/COVID-19

https://journals.plos.org/ploscompbiol/article?id=10.1371/journal.pcbi.1005697#sec020

https://www.statcan.gc.ca/eng/start

https://data.census.gov/cedsci/table?q=Florida&tid=ACSST1Y2019.S0101&hidePreview=false

https://population.un.org/wpp/Download/Standard/Population/

https://www.covid19.healthdata.org

https://www.statista.com/statistics/1114375/wearing-a-face-mask-outside-in-european-countries/

## Declarations

## Acknowledgements

Not applicable.

## Abbreviations

Not applicable.

## Availability of data and materials

The data used and/or analyzed during the current study are derived from public domain resources. The datasets are available in the [COVID-19 Community Mobility Reports] repository, https://www.google.com/covid19/mobility/, the [COVID-19 Data Repository by the Center for Systems Science and Engineering (CSSE) at Johns Hopkins University], https://github.com/CSSEGISandData/COVID-19. We used the [Prem et al. contact data], https://journals.plos.org/ploscompbiol/article?id=10.1371/journal.pcbi.1005697sec020. Population data were obtained from Stats Canada https://www.statcan.gc.ca/eng/start, US Census data https://data.census.gov/cedsci/table?q=Floridatid=ACSST1Y2019.S0101hidePreview=false, and https://population.un.org/wpp/Download/Standard/Population/ for other countries. The data related to mask wearing compliance are available on covid19.healthdata.org and https://www.statista.com/statistics/1114375/wearing-a-face-mask-outside-in-european-countries/.

## Ethics approval and consent to participate

This study does not require ethical approval. All data used or analyzed in this study are freely available in the public domain, and all methods are obtained directly from cited researchers. There is no direct interaction between the researchers and any individual or group in this study.

## Consent for publication

Not applicable.

## Authors’ contributions

All authors contributed equally to the design and implementation of the research, analysis of the results, and writing of the manuscript.

## 5 Appendix

### 5.1 Next generation matrix and reduced Jacobian

The Jacobian matrix near the disease-free equilibrium (DFE, which consists of *S*(0) = *N* and *I*(0) = 0) for the system of equations (2) is:

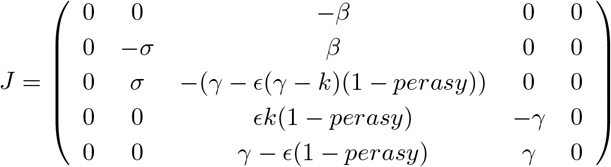

Using the next generation matrix method around the DFE ([34]) we compute *R*_0_ as the largest eigenvalue of the matrix *FV* ^−1^ and we obtain its closed form expression:

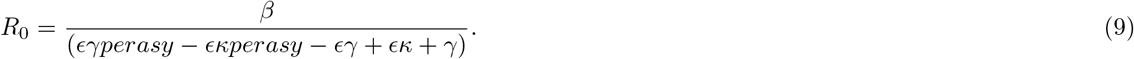

Further, using [55], we can compute the eigenvalues of the reduced Jacobian above and find that there is one positive eigenvalue (responsible for the growth near the DFE) which can be derived in closed form:

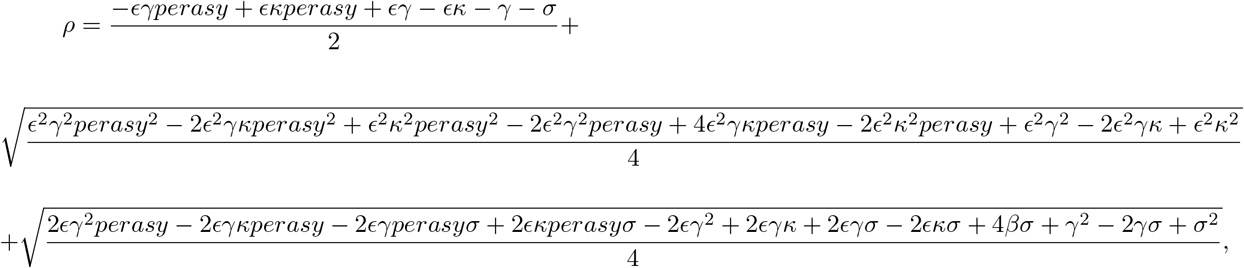

which in turn can be solved for an expression of *β* as a function of the growth factor *ρ* near the DFE:

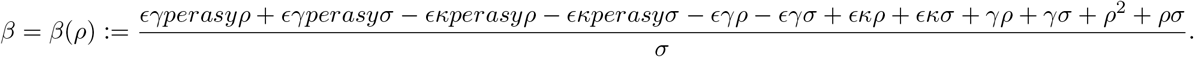

Finally we can estimate *R*_0_ as a function of the growth factor near the DFE in each region using 9 as:

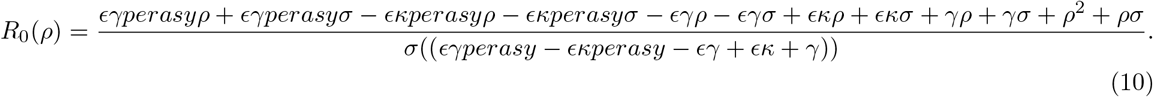

### 5.2 Mask Efficacy

First several months of the pandemic, there was considerable debate on the effect of face masks on limiting the spread of the COVID-19 pandemic and whether to recommend the general public to use a face mask. Later, articles, scientific reports, and data proved the impact of face masks in altering the outcomes of peak hospitalization [5]. Notably, face masks are found to be useful in both preventing asymptomatic transmission and illness in healthy persons. We adapt our previously developed SEIRL model [56] for transmission of COVID-19 with the impact of public use of face masks. Moreover, varying efficacy and compliance of masks have an impact on the transmission dynamics and control of the COVID-19 pandemic [4].

A review [57] of observational studies estimates that surgical and comparable cloth masks are 67% effective in protecting the wearer. Some reports show that even a cotton T-shirt can block half of the inhaled aerosols and almost 80% of exhaled aerosols measuring 2*μm* across e.g. unpublished work by Linsey Marr, an environmental engineer at Virginia Tech in Blacksburg. We consider 50% efficacy in our model.

### 5.3 Compliance with Mask

The proportion of a population wearing face masks differs across countries/regions based on social norms, political reasons, the consequences of non-compliance e.g., fines. The results from a study surveying are different for example:

- The Institute for Health Metrics and Evaluation (IHME), a global health research center at the University of Washington [51] is reported the percentage of mask use in Italy was between 63% to 93% from September 1st till December 31, 2020, Sweden 1-7%, Saudi Arabia 73-76%, Ontario 75-85%, Florida 66-70%, Romania 63-86%, Ghana 50-36%, South Africa 80-81%, Indonesia 74-76%, Nepal 64-63%, Brazil 68-59%, Argentina 89-83%.
- According to data from the Institute for Health Metrics and Evaluation at the University of Washington in Seattle, mask use has held steady around 50% since late July in the United States. It was predicted to increase to 95% as of 23 September. (see [51]) Whereas, a survey from Gallup [58] shows 72% of U.S. adults say they either always wear a face mask or wear one often when going to public places.
- Percentage of people who worn a face mask outside their home always is reported 93.9% in Italy and 12.1% in Sweden 12.1% by YouGov; Imperial College London [52].

We adapt our SEIRL model with the compliance of mask-wearing value denoted as *compl* in table 1 for each region.

### 5.4 Deriving average contact rates

In contact transmissible diseases e.g. COVID-19, contact rate has an important role in epidemic models. Prem et al. [21] provide data-driven contact matrices in the home, work, school, and other locations for 152 countries of the world. We amalgamated the average contact rate in home, work, and other locations, then we computed the weighted average of a given projected contact matrix in a region with the corresponding proportions of 5-year age population groups in 2020 to determine one single average contact rate.

## Footnotes

[1] We chose to concentrate on the year 2020 specifically because there were no preventative or antiviral treatments known against this virus at that time, and countries had to rely on some combination of NPI’s to fight its spread.

[2] The last column in Table 2 highlights the number of weeks that `dpseg` is assigning for the steepest slope. The weeks are numbered from the first week with positive cases in each location, so near disease-free.

